# Caught in the data quality trap: A case study from the evaluation of a new digital technology supporting routine health data collection in Southern Tanzania

**DOI:** 10.1101/2023.04.12.23288456

**Authors:** Regine Unkels, Aziz Ahmad, Fatuma Manzi, Asha Kasembe, Ntuli A. Kapologwe, Rustam Nabiev, Maria Berndtsson, Atsumi Hirose, Claudia Hanson

## Abstract

**Background:** Health service data from Health Management Information Systems is important for decision-making at all health system levels. Data quality issues in low-and-middle-income countries hamper data use however. *Smart Paper Technology*, a novel digital-hybrid technology, was designed to overcome quality challenges through automated digitization. Here we assessed the impact of the novel system on data quality dimensions, metrics and indicators as proposed by the World Health Organization’s *Data Quality Review Toolkit*.

**Methods:** This cross-sectional study was conducted between November 2019 and October 2020 in 13 health facilities sampled from 33 facilities of one district in rural Tanzania, where we implemented *Smart Paper Technology*. We assessed the technology’s data quality for maternal health care against the standard *District Health Information System-2* applied in Tanzania.

**Results:** *Smart Paper Technology* performed slightly better than the *District Health Information System-2* regarding *consistency between related indicators* and *outliers*. We found <10% difference between related indicators for 62% of the facilities for the new system versus 38% for the standard system in the reference year.

*Smart Paper Technology* was inferior to *District Health Information System-2* data in terms of *completeness. We observed that data on 1*^*st*^ *antenatal care visits* were complete ⍰ 90% in only 76% of facilities for the new system against 92% for the standard system. For the indicator *internal consistency over time* 73%, 59% and 45% of client numbers for antenatal, labour and postnatal care recorded in the standard system were documented in the new system. *Smart Paper Technology* forms were submitted in 83% of the months for all service areas.

**Conclusion:** Our results suggest that not all client encounters were documented in *Smart Paper Technology*, affecting data completeness and partly consistency. The novel system was unable to leverage opportunities from automated processes because primary documentation was poor. Low buy-in of policymakers and lack of internal quality assurance may have affected data quality of the new system. We emphasize the importance of including policymakers in evaluation planning to co-design a data quality monitoring system and to agree on a realistic way to ensure reporting of routine health data to national level.

## BACKGROUND

Well-functioning health management information systems (HMIS) are an important part of a country’s health system particularly for sub-national level evidence-informed decisions on health service delivery. HMIS data in many low-and-middle-income-countries (LMIC) including Tanzania are collected by health care providers (HCP) in multiple paper-based facility registers, then summarized in facility summary reports. Since 2013 this summary data is then manually digitized in the electronic HMIS data platform called *District Health Information System-2* (DHIS2) (1-3) (supplementary table S1).

Research from several settings indicated HMIS data quality issues in terms of completeness and consistency (4-7). The reported shortcomings may be related to the fact that data is partly processed manually, first documented on typically paper-based registers and then tallied and summarized. This process is prone to calculation and transfer errors (8, 9) raising the need to innovate systems overcoming these errors.

*Smart Paper Technology* (SPT) is one approach to digitalize data processing and may improve data quality through automatization (10). SPT is an innovative digital-hybrid system using scannable paper forms matching facility register content (supplementary file S1) and was introduced under a pilot project in all 33 health facilities providing maternity care in Tandahimba district in a phased approach between June 2019 and July 2020. Our pilot project implemented SPT to process antenatal care (ANC), labour care (LC) and postnatal care (PNC) data (figure 1 below).

**Figure 1:**
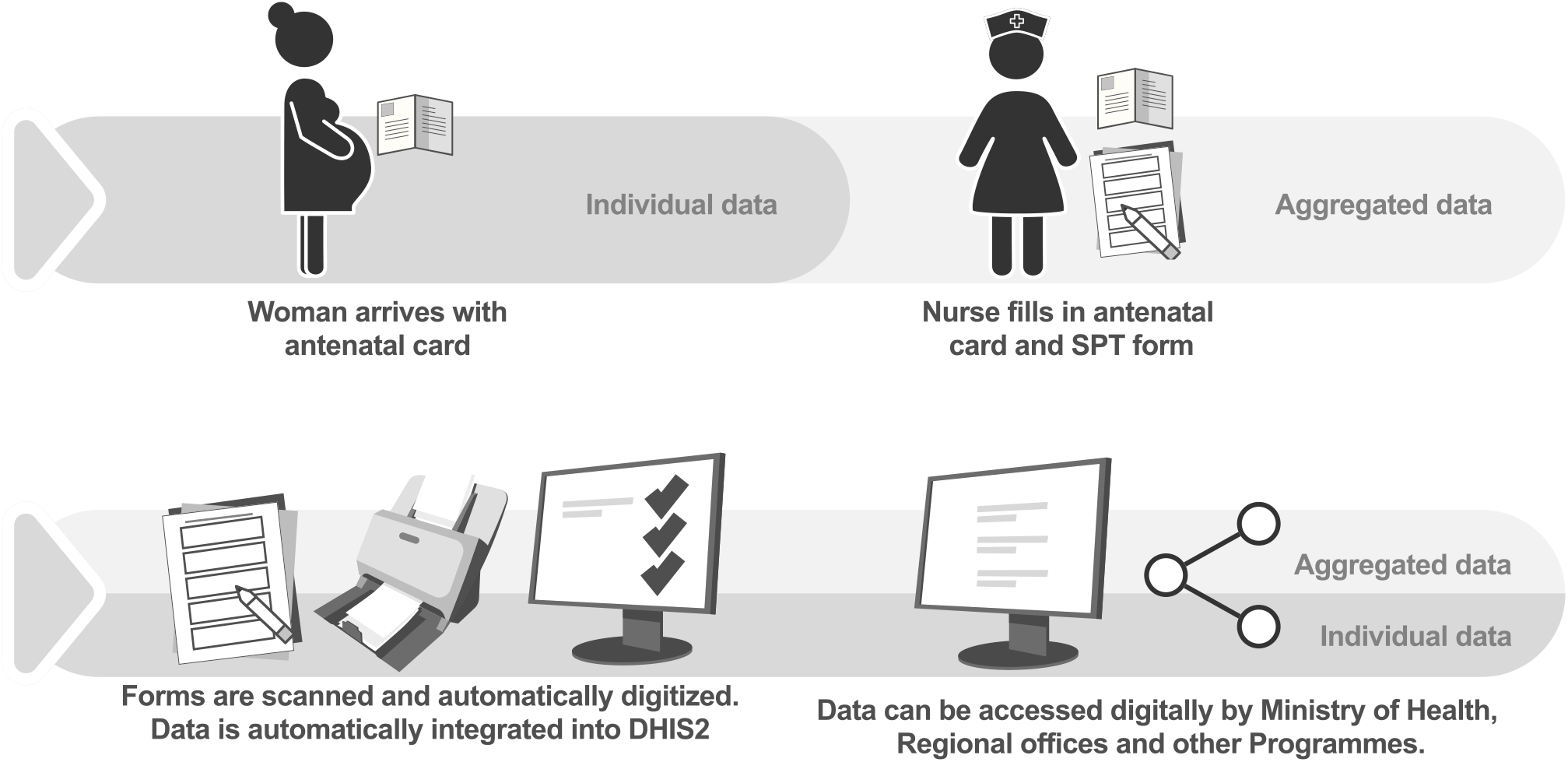
Smart paper technology.

Contrary to facility registers, filling SPT forms requires only a tick instead of written text. Each woman receives a unique identifier at registration which is subsequently used on all forms during ANC, LC and PNC. Forms are automatically digitized during scanning and electronic SPT summary reports are created by special software (figure 2 below). The system uses the identifier to generate individualized data throughout data processing (figure 1).

**Figure 2:**
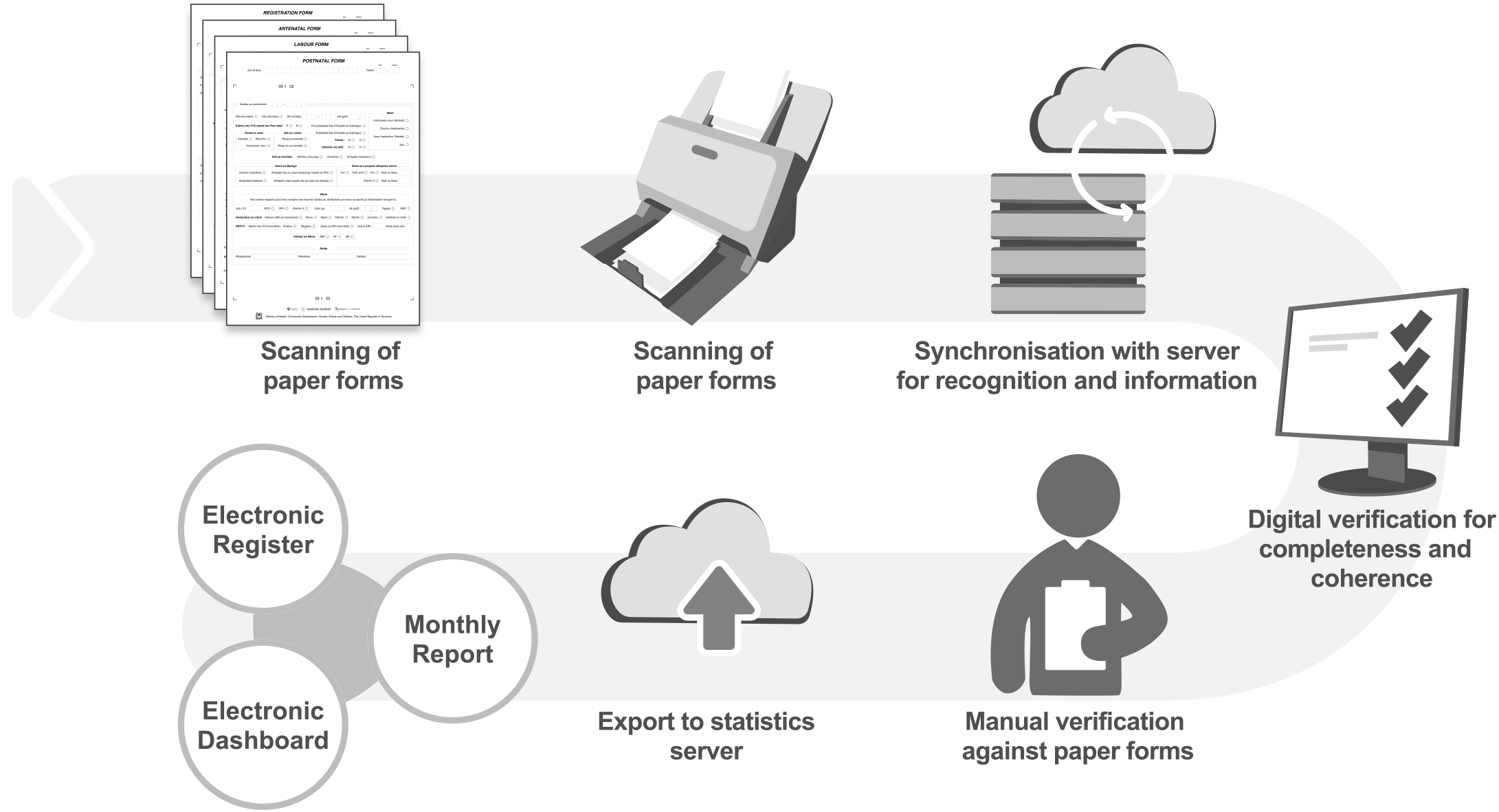
Data processing with smart paper technology. Legend: Verification flagged information that was either incorrectly entered or recognised. Incoherent information such as unusually low or high values for client age was identified and flagged by the software. Information that was expected to always be filled in, such as place and mode of delivery, was flagged if no data was recognised. Manual verification included including quality assurance, where automated verification was checked by a research team member (AA) who provided feedback to verification officers.

SPT can potentially improve HMIS data quality through simplified primary data entry and automated digitization. Studies reported good SPT data quality for vaccination services in The Gambia and Uganda (11, 12). Our previous findings from the process evaluation of this novel system suggest that SPT can be embedded into existing maternal health care provision, is acceptable and potentially generates time-savings for HCPs (13).

The World Health Organization published guidance on standardized data quality assessments in 2017 to facilitate regular national and sub-national reviews of HMIS data quality in LMICs. This Data Quality (DQR) Toolkit defines four quality dimensions, i) *data completeness and timeliness*, ii) *internal consistency*, iii) *external comparison* and iv) *external consistency with population data*, each with a set of metrics and indicators (14) and is now increasingly used for data quality assessments in various LMIC countries including Tanzania allowing comparison between countries (4, 5, 15, 16).

In this study we applied the WHO’s DQR toolkit to evaluate SPT data quality for maternal care services in terms of i) *completeness and timeliness* and ii) *internal consistency* to further inform understanding of its scale-up potential. We assessed data quality dimensions for key indicators of ANC, LC and PNC services at all the three levels of health care in Tandahimba district in rural Tanzania.

## METHODS

### Design

We conducted a cross-sectional study on the quality of SPT data routinely collected by 13 health facilities for maternity care services in Tandahimba district in Southern Tanzania between November 2019 and October 2020. This study was part of a process evaluation of SPT implementation in all 33 health facilities providing maternal care in the district (13). Results are reported using the *Strengthening the Reporting of Observational Studies in Epidemiology* (STROBE) statement (17).

### Setting

Tandahimba is a typical rural district in terms of health care delivery and reported challenges with HMIS data quality (5). One district hospital, two health centers and 10 dispensaries were included. All the three facility types provided ANC, LC and PNC services. Dispensaries typically offer basic maternity care for low-risk mothers. Clients requiring advanced services are referred to health centers, or further to a district hospital.

**SPT data processing** included the completion of paper forms for each client contact at health facilities and their transfer to the district level for scanning, synchronisation with a server and automated recognition and verification (figure 2). Monthly electronic summary reports were displayed on the SPT dashboard on the 6^th^ of each month (supplementary figure S1). This date was later changed to 10^th^ of each month to synchronize with transport of facility summary report forms to district headquarters for DHIS2 data entry.

**DHIS2 data processing** followed the described path of documentation in i) facility registers, and ii) summarized monthly facility summary reports which were then transported to the district headquarters at the latest on the 10th of each month for manual data verification and digitization. Physical visits to health facilities for data verification were often necessary when obviously implausible data was identified during DHIS2 data entry. Verification and DHIS2 entry could take up to two weeks, typically involving several managers, until data was available on the DHIS2 dashboard by the 17^th^ of each month (18). The Ministry of Health had requested that HCP enter client data in both facility registers and SPT forms throughout the evaluation period to maintain routine reporting to national level.

### Sampling procedure and sample size

We chose the 13 study sites for logistic reasons: i) Staff were already trained on SPT use in November 2019, ii) and had completed one year of data collection when the SPT project ended to include likely seasonal variations. No sample size calculation was performed.

### Outcome variables

We adapted selected quality metrics and indicators from the DQR Toolkit for the evaluation of SPT data quality reflecting service provision data from the continuum of maternity care. **Table 1** provides a detailed description and definitions of adapted data quality dimensions, metrics and indicators with their respective benchmarks.

**Table 1.**
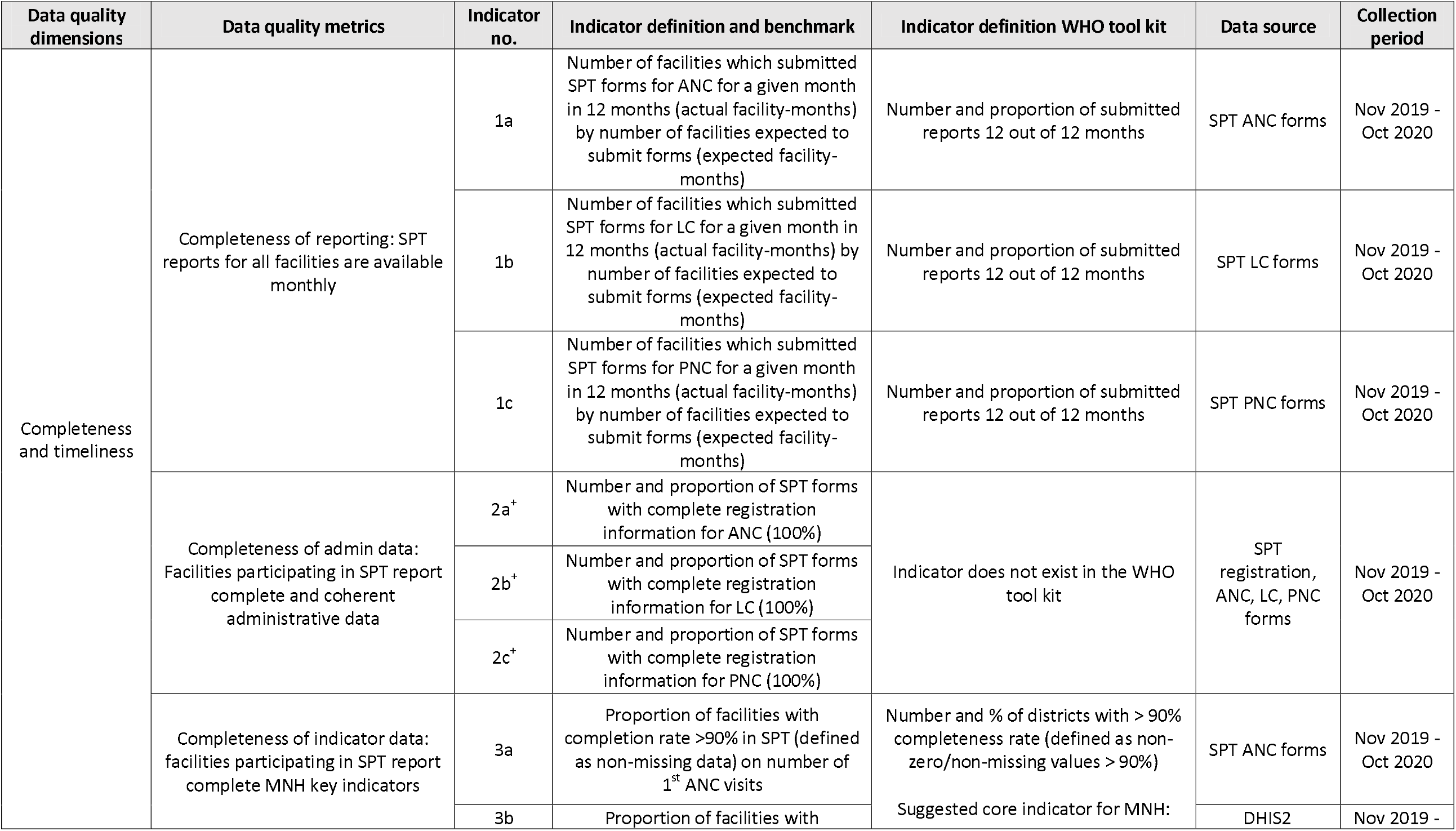
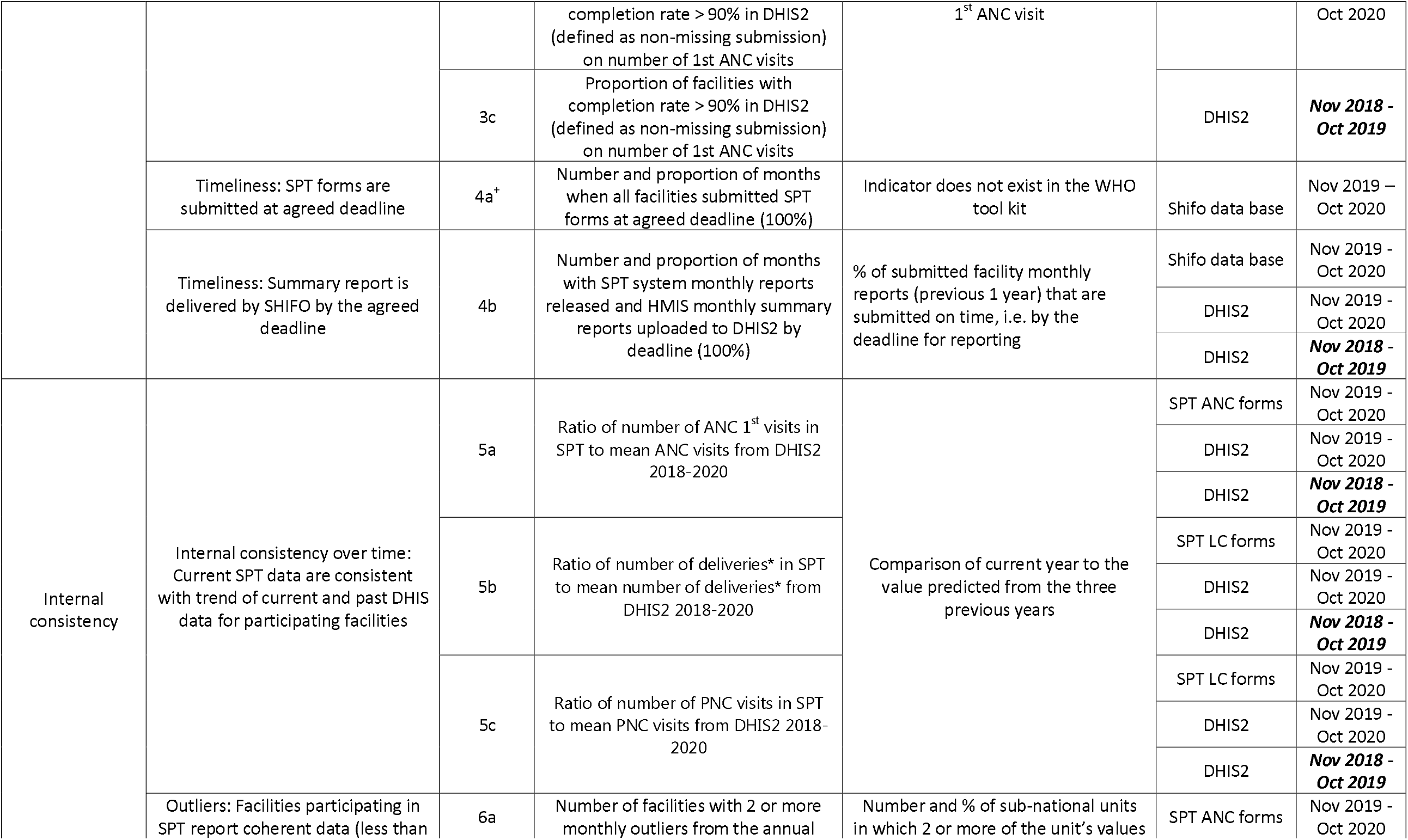
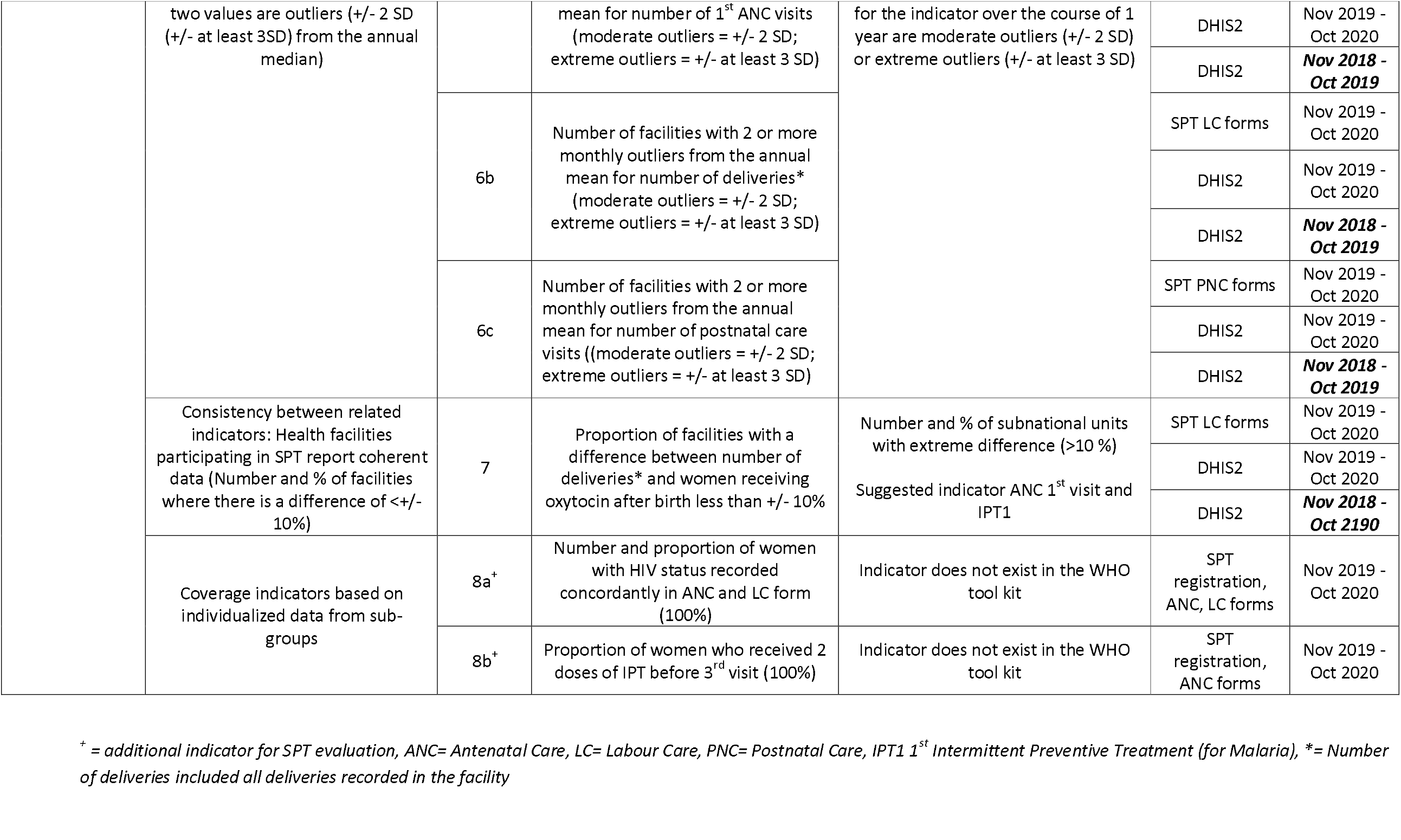
Metrics and indicators for data quality dimensions of completeness, timeliness and consistency of SPT and DHIS2.

Our main outcome variables included the following two quality dimensions, metrics and indicators: i) *completeness and timeliness* of data and ii) *internal consistency*, using DHIS2 data as comparison where possible (table 1). Digital DHIS2 data was chosen over facility register data because the first is used for national and sub-national planning and performance monitoring, while registers remain in facilities without further use.

#### Completeness and timeliness

We developed two *additional indicators on completeness and timeliness of reporting* to include SPT form submission (indicator 2 and 4a in table 1) which has no equivalent in DHIS2.

We defined completeness as i) SPT form submission from ANC, LC and PNC each month for the entire evaluation period (12 months) (**indicator 1**), ii) continuous application of the SPT unique identifier (**indicator 2**) and iii) complete reporting on first antenatal care visits each month without missing monthly values (**indicator 3**).

Timeliness was defined as i) timely receipt of SPT forms on agreed date at district headquarters (before 6^th^, later 10^th^ of each month) (**4a**), ii) timely release of electronic summary reports on the SPT dashboard (6^th^, later 10^th^ of each month) and on DHIS2 (17^th^ of each month) (**indicator 4b**).

#### Internal consistency

A total of two *additional indicators* was developed to evaluate SPT’s potential to generate individualized data based on the unique identifier, allowing the computation of coverage data for client sub-groups, which is impossible to obtain from DHIS2 (indicators 8a, and b, table 1).

We included three data quality metrics for selected maternal health care indicators: i) data consistency over time (**indicator 5**), ii) presence of outliers (**indicator 6**) and iii) consistency between related indicators (**indicators 7 and 8**). *Moderate outliers* were defined as monthly values diverting from the annual mean numbers of ANC 1^st^ visits, deliveries or PNC visits in each facility by two standard deviations (SD) in any direction. *Extreme outliers* were defined as monthly values diverting by three standard deviations or more (14, 19). *Related indicators* were defined as indicators with a predictable relationship (14), in our case, i) the number of deliveries and the number of women who received oxytocin after delivery (**indicator 7**), ii) number of women with concordant HIV-status in ANC and LC forms (**indicator 8a**) and iii) the number of women receiving their 2^nd^ intermittent preventive treatment against malaria during their 2^nd^ ANC visit (**indicator 8b**).

### Data sources

We used routine data processed in SPT and DHIS2 on ANC, LC and PNC. We included DHIS2 data from two time periods, November 2018 - October 2019 and November 2019 - October 2020. We refer to the latter DHIS2 data as the reference year. Data from the previous year were included to mitigate effects of interdependency between SPT and DHIS2 during the reference year due to duplicated data entry (20). We could not identify any reference data source for additional indicator 8 because other available databases mostly used aggregated DHIS2 data.

### Data collection

To evaluate timeliness, we extracted i) scanning dates logged in the SPT system, ii) dates of electronic SPT summary report release and iii) upload dates of facility summary reports into DHIS2. Data on completeness and internal consistency was downloaded from SPT and DHIS2 databases by the research team in February 2021.

### Data processing and analysis

Descriptive statistics with simple frequencies, ratios and data trends were generated in STATA 16 (StataCorp LLC, Texas, USA) for monthly evaluation time points. No confidence intervals were calculated due to the low number of included health facilities.

### Ethical considerations

All methods were carried out in accordance with relevant guidelines and regulations. <u>E</u>thical clearance was obtained for the overall SPT evaluation from the *Institutional Ethical Committee of Ifakara Health Institute* (IHI/RB/No.20 -2018) and *National Institute of Medical Research* (NIMR/HQ/R.8a/Vol.IX/3018) in Tanzania and from the *Ethics Review Board of the Commune of Stockholm* (2019-04022 Gk), Sweden. Permission to use the two data bases for our study was granted by the Ministry of Health, Community Development, Gender, Elderly and Children, the President’s Office, Regional Administration and Local Governments and the respective Regional Administrative Secretaries and Hospital Authorities during stakeholder consultations. The need for individual consent was thus deemed unnecessary according to national regulations and this approach was approved by the IRB of Ifakara Health Institute and National Institute of Medical Research. No identifiable variables such as names of individuals were collected during this study. Names of health facilities involved in the study were not used for reporting. No administrative permissions apart from the above mentioned were required to access the raw data used in our study.

## RESULTS

A total of 13,904 individual ANC forms, 3,596 LC forms and 3,895 PNC forms were processed in the SPT system between November 2019 and October 2020.

### Completeness and timeliness

All 13 health facilities were expected to submit SPT data each month over the 12 months evaluation period, for the total of 468 facility-months for all three service areas (156 each for ANC, LC, and PNC). SPT forms were submitted for 389 actual facility-months (83% of the expected number), with ANC submission being most complete (150 (96%)), followed by LC (128 (82%)) and PNC with the lowest completeness of form submission (111 (71%)) (table 2) (**indicator 1**).

**Table 2:**
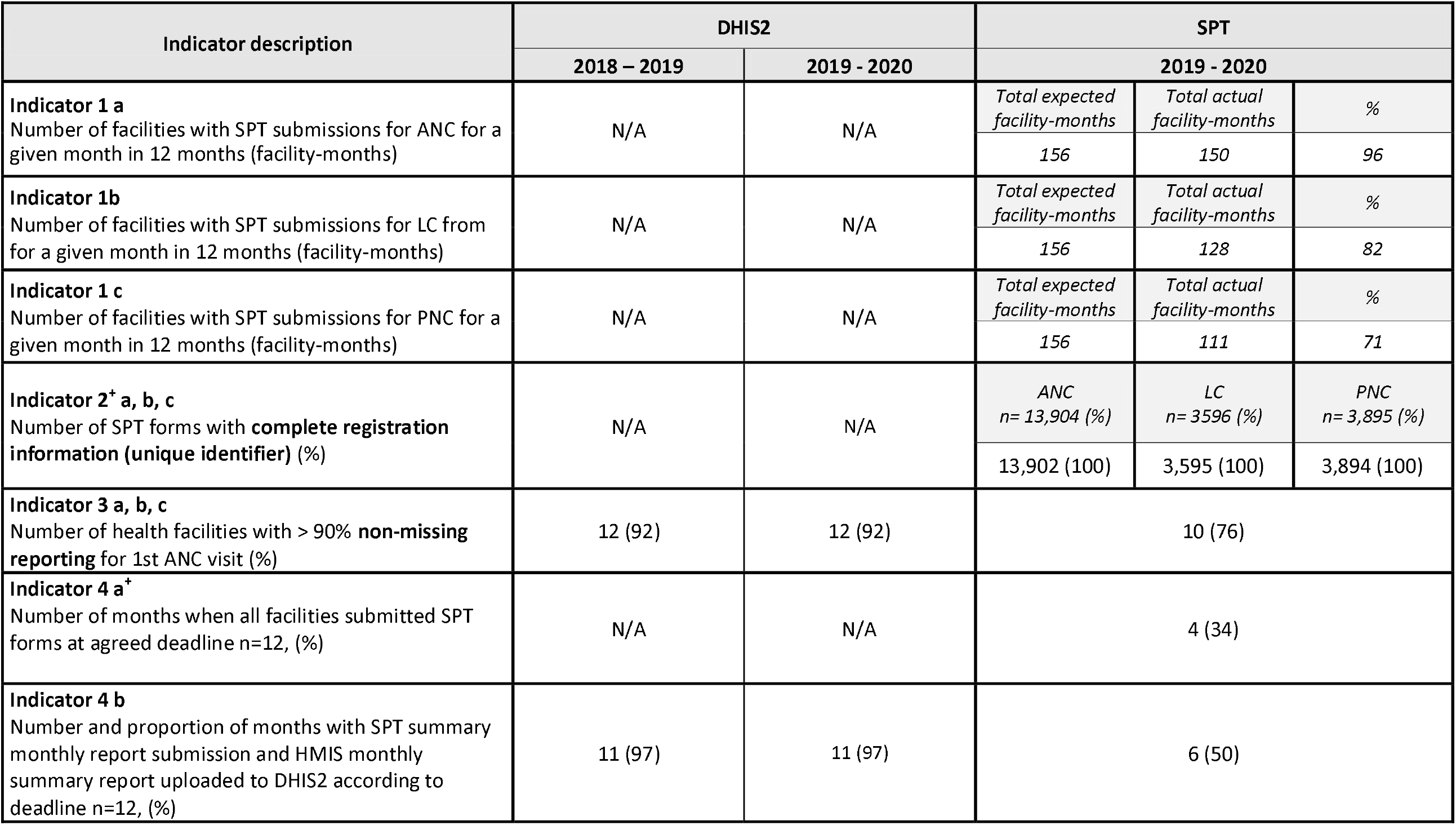
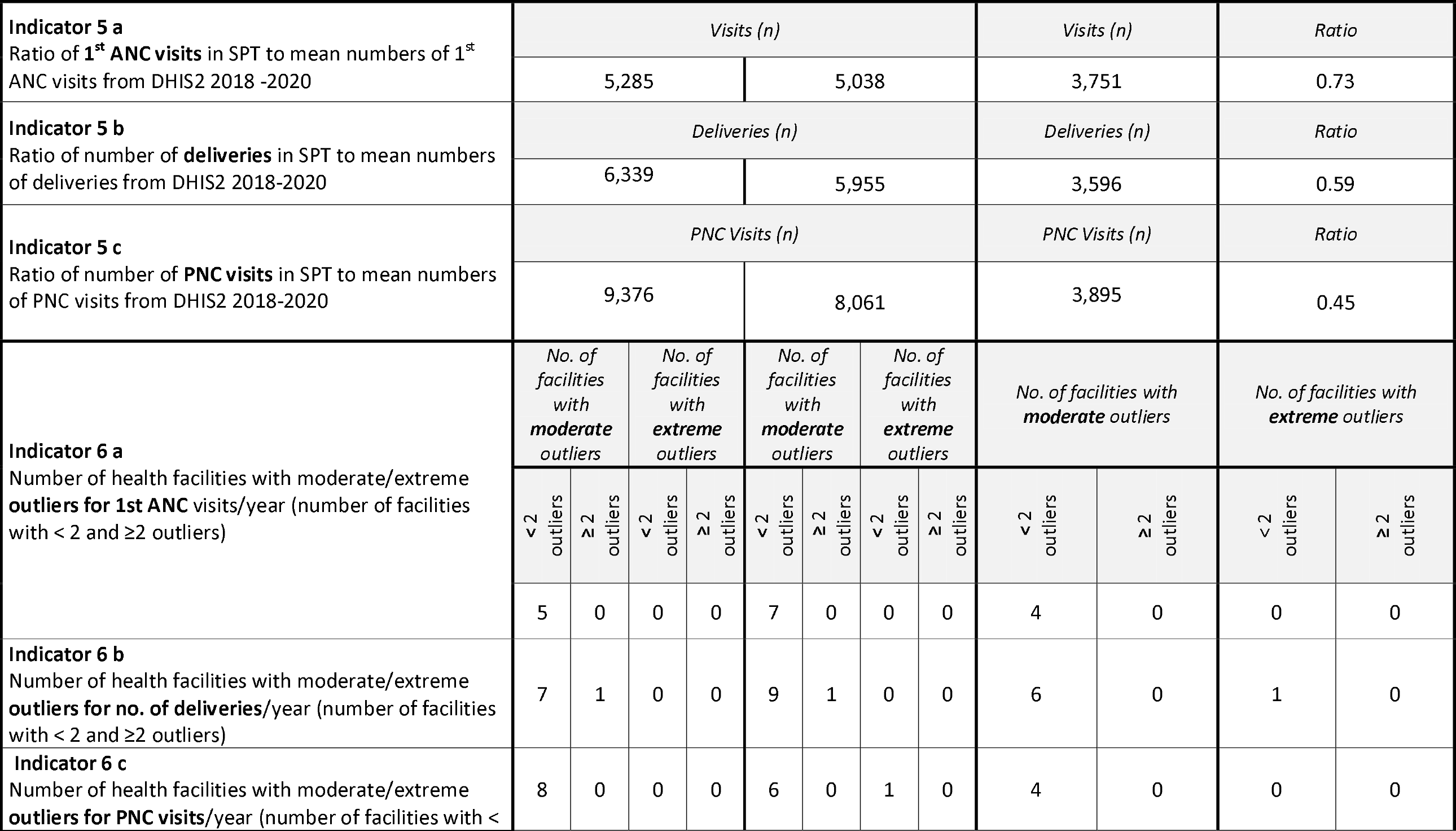
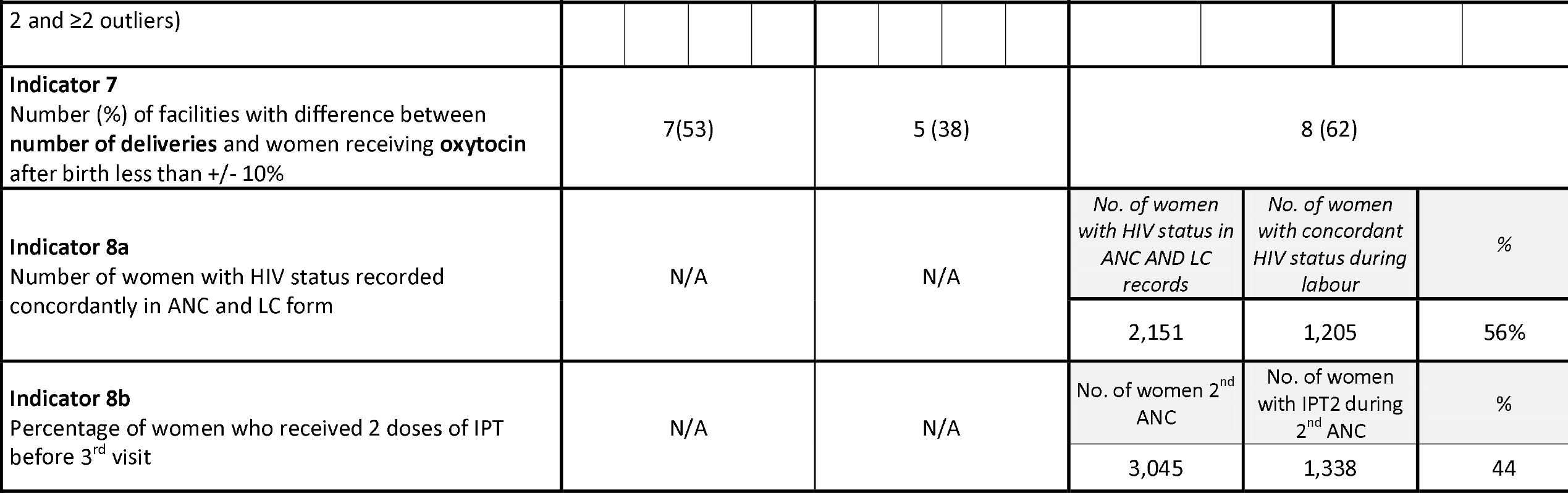
Data completeness, timeliness and consistency of SPT and DHIS2 (facilities n=13, if not otherwise stated)

Registration information completeness, measured as the continuous use of the unique identifier, was found to be highly complete at 100% of the forms throughout all three service areas (table 2) (**indicator 2**^+^).

SPT data on the number of 1^st^ ANC visits were > 90% complete (without missing data on 1^st^ visits each month) for 10 facilities out of 13 (76%) during the 12 months evaluation period as compared to 12/13 (92%) for DHIS2 2018-20 (table 2). One dispensary submitted complete SPT data on numbers of 1^st^ ANC visits for 6 months only, while submitting complete data for 11 months for DHIS2 2018-19 and for 12 months for DHIS2 2019/20. Without this facility, overall SPT completeness would have been 83% (supplementary table S2) (**indicator 3**).

Timely submission *of SPT forms* on the agreed date (6^th^ of each month) from all facilities to the scanning station was achieved in four out of 12 months (34%) (**indicator 4a**^+^). Timely dashboard display of electronic *SPT summary reports* was achieved in six out of 12 months (50%). The standard electronic DHIS summary reports were displayed timely for 11 out of 12 months (97%) (table 2) (**indicator 4b**).

### Internal consistency

The mean number of ANC 1^st^ visits, deliveries and PNC visits reported through DHIS2 were 5,285 (ANC 1), 6,339 (number of deliveries) and 9,376 (PNC) in 2018/19 and 5,038 (ANC 1), 5,955 (Deliveries) and 8,061 (PNC) in 2019/20 respectively (table 2). SPT reported 3,751 ANC 1^st^ visits, 3,596 deliveries and 3,895 PNC visits, indicating a ratio of SPT numbers to the mean numbers from both DHIS2 reports of 0.73 (ANC 1), 0.59 (number of deliveries) and 0.45 (PNC). Documented monthly SPT client numbers as compared to the DHIS2 reference year were lower for **each month** for all three service areas (supplementary figure S2) (**Indicator 5**).

Both SPT and DHIS2 showed few monthly outliers overall from the annual mean of number of ANC visits, deliveries or PNC visits in each facility. But while the SPT system displayed no facility with two or more outliers, both DHIS2 data sets included two or more facilities with outliers for LC. One facility had an extreme outlier (defined as +/- at least 3 SD) in SPT for LC, and one facility had an extreme outlier in the reference DHIS2 for PNC (table 2) (**Indicator 6**).

The number of deliveries and number of women receiving oxytocin after birth, which should be almost equal, showed higher consistency for SPT than for both DHIS2 data sets. An acceptable difference of less than 10% between both indicators was recorded for 62% of facilities in SPT, compared to 38% for DHIS2 2019/20 and for 53% for DHIS2 2018/19 (table 2). The mean difference for all 13 health facilities for SPT was 9% compared to 10% for DHIS2 2018/19 and 12% for DHIS2 2019/20 (supplementary table S3) (**Indicator 7**).

We included 2,151 women in the analysis of **indicator 8a**^+^ where information from SPT ANC and LC forms about HIV-status could be linked through the unique identifier. We found that the documented HIV-status was consistent in linked forms for 1,205 (56%) of these women (table 2).

We included ANC forms of 3,045 women where information on service provision and number of ANC visit could be linked through the unique identifier in the analysis of **indicator 8b**^+^. We found that for 1,338 (44%) of these women, intermittent preventive treatment (IPT) for malaria was correctly reported as the second dose during their 2^nd^ ANC visit (table 2).

## DISCUSSION

Our study evaluated SPT data quality for selected maternal health indicators using DHIS2 data as a benchmark. We observed a mixed data quality pattern where completeness of submitting the SPT papers to the scanning station was one important problem together with failure to document all client encounters in the SPT system. In contrast, the unique SPT identifier, important for internal consistency and the generation of cohort data, was used consistently in 100% across all three data sets of ANC, LC and PNC. We report that the new SPT system showed slightly more consistent data for *related indicators* for 62% facilities with an acceptable <10% difference between number of deliveries and oxytocin given at birth, against DHIS2 with 38% (2019-2020) and 53% respectively (2018-2019). Internal consistency of SPT data was also slightly better for the *presence of outliers* where fewer facilities reported moderate outliers for the SPT system.

In contrast, DHIS2 showed better data quality in terms of completeness, timeliness and for consistent data trends over time: While complete submission of specific information about 1^st^ ANC visits for the entire evaluation period was found in 92% in DHIS2, SPT data only achieved this n 76%. Timely submission of SPT forms from all facilities to the district capital was only found in 34% of the months. Consequently, electronic SPT summary reports were available on the dashboard in only 50% of the months on time compared to 97% availability of DHIS2 electronic summary reports.

Our findings on completeness are in contrast with other studies assessing SPT data quality for vaccination services against HMIS data from the previous year (11). It is to note that in The Gambia SPT replaced facility registers, and mandatory reporting of routine data was transferred to SPT forms. In contrast, in Tanzania, the ministry of health, the owner of DHIS2, required duplicate data collection for this pilot project.

While we accept that the low completeness is clearly disfavoring the SPT system, operational and practical reasons may explain this. Firstly, HCPs may have prioritized data entry for DHIS2 as the formal public system. Our research project had no mandate to enforce documentation using SPT forms, instead introduction of SPT led to duplicated documentation. We previously reported on qualitative findings from the overall SPT evaluation where HCPs and their managers described how this may have contributed to incompleteness of SPT and HMIS data (13). We noted from our current data that DHIS2 for 2018-2019 showed slightly better *internal consistency* than in the reference year after SPT introduction (2019-2020) (indicators 6 and 7, table 2). This finding may suggest that the reference DHIS2 could have also been incomplete to a certain extent. Other studies on HMIS data quality support this assumption (5, 6, 8, 13). Furthermore, HMIS research using the *DQR Toolkit* has confirmed upward trends of maternal coverage indicators over three years due to population increase as described in the *DQR Toolkit* (4, 14, 19). In contrast we noted stable client numbers for ANC 1^st^ visit, deliveries or PNC within DHIS2 during the two years included (indicator 5) (supplementary figure S2). Although a slightly reduced maternal health service provision was reported for Tanzania during the COVID-19 pandemic (−2.6 % for ANC 1^st^ visit, -6.8% for labour care) (21), it is difficult to attribute the trend we observed to these circumstances alone.

Our findings reported here, together with results from our process evaluation (13) suggest, that institutionalization of SPT was not achieved, possibly due to low managerial buy-in although HCPs saw the technology’s benefits for their work. We argue that low data quality of SPT may have contributed to this situation and at the same time, lack of buy-in and institutionalization perpetuated duplicate data entry and thus low data quality (figure 3 below).

**Figure 3:**
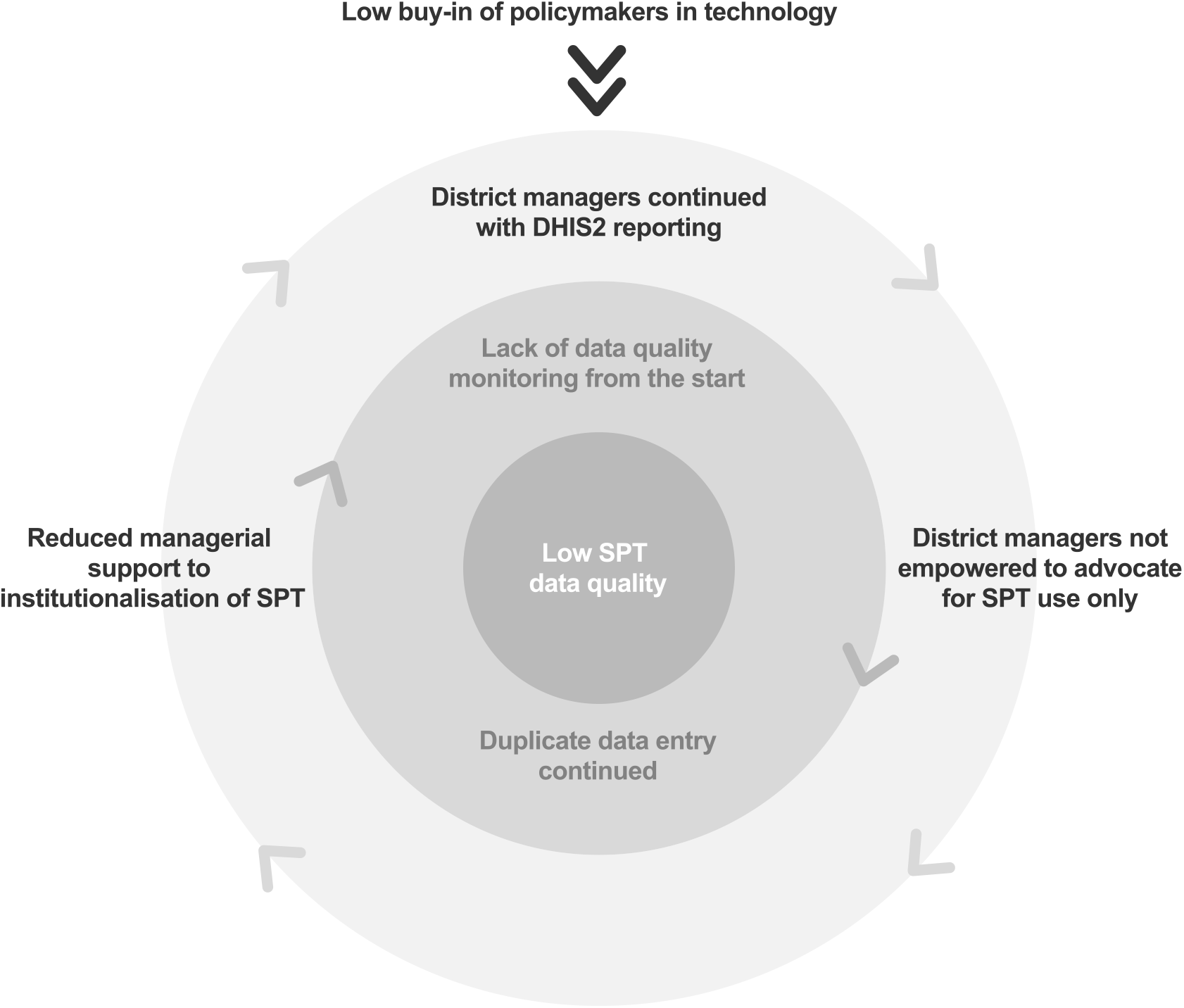
Duplicate data entry perpetuated low data quality and low trust in SPT data.

Secondly, maternal health care is more complex than child vaccination services, where SPT was initially assessed (11, 12). The latter deliver and document vaccinations within one service area during four outpatient visits. Maternal health services in contrast, include three service departments along the continuum of antenatal, labour and postnatal care. Continuous documentation is thus more complex, also because at hospital level, ANC and partly PNC are outpatient departments, while labour care is provided as an inpatient service. Unique identifiers may thus not have been immediately available from other departments for each client and HCPs may not always have taken the trouble to trace it, but simply not complete the SPT form, especially at night when outpatient departments are closed. This complexity may have impacted negatively on SPT data quality (12, 13).

Thirdly, contextual factors related to health system challenges may have also affected SPT data processes and eventually data quality. Evidence suggests a multitude of, mostly contextual, underlying causes for data incompleteness and inconsistency for HMIS and it is likely that these have also influenced SPT data quality (8, 16, 22). Previous studies support this hypothesis, describing the effects of i) documentation supply stockout and ii) lack of supervision and human resources and iii) an organizational culture that valued summary data over primary data and completeness over correctness (13, 23): Timeliness of SPT form submission mainly depended on transportation from facilities to district headquarters. The deadline for electronic summary report display on the SPT dashboard was consequently changed to allow synchronized transportation of SPT forms and facility summary reports. Our results related to IPT provision during ANC (indicator 8b, table 2) may suggest that not only incomplete documentation, but also incomplete service provision could have contributed to the low data consistency, e.g. caused by stock out of sulphadoxine-pyrimethamine for IPT (24, 25).

The described barriers may have prevented users from fully reaping SPT benefits from digitalization for data quality. We hypothesize that the quality of primary collection of routine health information is not only dependent on the technology (e,g, SPT or HMIS) but even more on contextual and individual factors which were likely to be similar for both systems. We also emphasize the importance of national sovereignty over data and data systems (26) as a contextual factor influencing stakeholder buy-in for digitalization projects.

We believe that our findings on SPT data quality are important despite the constraints described above. The interdependency of data quality and context allowed us to note key constraints for the implementation and evaluation of a new digital technology which would have been otherwise difficult to detect during implementation. These quantitative findings triangulate other findings from the process evaluation of SPT introduction (13), but add another dimension to evaluations of the so called “*eco-system of digitalization*” (27, 28). They also put into perspective other studies describing the validation of international indicators based on routine health data (29): Due to the mutual existence of several data processing systems in Tanzania (23), duplicate data entry is rather the norm than an exception, also for HMIS data, with the consequences described.

### Methodological considerations and limitations

We note the limitations of our methodology. Firstly, SPT and DHIS2 from the reference year were interdependent, containing the same primary data, entered by the same users in the same context (20). We partially mitigated potential effects by including DHIS2 data from the year before SPT introduction in the health facilities (11). Secondly, DHIS2 data is the foundation of the Government’s resource planning, and thus advocating SPT use only was difficult.

Thirdly, we were unable to assess the *DQR Toolki*t’s quality dimensions of *external consistency with different data sets and with population data*. Population data for *Demographic and Health Surveys* is aggregated at regional level (30), which did not allow for comparison with our data from 13 health facilities (14). Other local registers also used DHIS2 data and could thus not be applied. External comparison, however, would have been important.

Definitions of HMIS data quality and its dimensions differ in the literature (6, 7, 29, 31-33), making standardized measurements and comparison between studies difficult. We therefore chose the *DQR Toolkit* to strengthen comparability of our results with other SPT or HMIS data quality assessments (4, 5, 11) but our experiences suggest that additional methods may be needed to address the key issues we identified.

## CONCLUSIONS

Our results suggest that SPT performed well in terms of internal consistency, but completeness was low. The main reason for the low quality was probably that this pilot project was unable to fully implement the system. In response, HCPs were requested to report data twice which understandably reduced their commitment resulting in low completeness. Our findings thus provide little information on the data quality that theoretically could be achieved by SPT but rather highlight that data quality issues result from inadequate implementation. Data quality should probably be seen as the outcome of operational and systemic factors rather than a specific attribute of technology processing health data.

We conclude that sustained stakeholder involvement is important during planning of evaluation studies on digital technology to support routine health data processing. In addition, we emphasize the need for a data quality monitoring strategy as an important implementation measure from the start of any project to ensure high quality evaluation data that can support decision-making on scale up.

## Supporting information

Figure S1 SPT dashboard

Figure S2 Indicator 5

File S1 sample SPT form

Table S1 Glossary of terms

Table S2 Indicator 3

Table S3 Indicator 7

## Data Availability

All relevant data is within the manuscript and the accompanying Supplementary files. The full SPT data set is stored on the institutional server of Ifakara Health Institute. In line with restrictions set by the Tanzanian ethics committees, access to the full data is only available to involved researchers on reasonable request. They will be asked to complete a data sharing agreement with the National Institute for Medical Research Ethics Committee and Ifakara Health Institute’s Institutional Review Board as per policy in Tanzania.
Data requests may be sent to the secretary of the independent Institutional Ethics Review Board associated to Ifakara Health Institute, Dr. Mwifadhi Mrisho via email to.

## List of Abbreviations

ANC: Antenatal Care
DHIS2: District Health Information System
DQR: Data Quality Review
HCPs: Health Care Providers
IPT: Intermittent Preventive Treatment
HMIS: Health Management Information System
LC: Labour Care
PNC: Postnatal Care
SPT: Smart Paper Technology

## Declarations

## Acknowledgements

We would like to thank the Tandahimba District Council and the President’s Office for Regional Administration and Local Government, PORALG for allowing us to conduct this study in their health facilities. We would especially like to thank Mrs Emily Kutandikila and Dr. Saidi Ngwalima for their support to the implementation of Smart Paper Technology in Tandahimba district council. We are grateful to Miss Sanni Kujala and Mr. Josef Przgodzicz from SHIFO for their support to the implementation of SPT.

