## Supplementary figures and images for "Caught in the data quality trap: A case study from the evaluation of a new digital technology supporting routine health data collection in Southern Tanzania"

### Figure S1 SPT dashboard

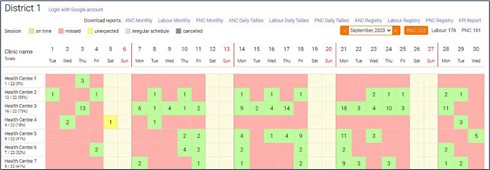
