## Supplementary material for "Caught in the data quality trap: A case study from the evaluation of a new digital technology supporting routine health data collection in Southern Tanzania": Figure S2 Indicator 5

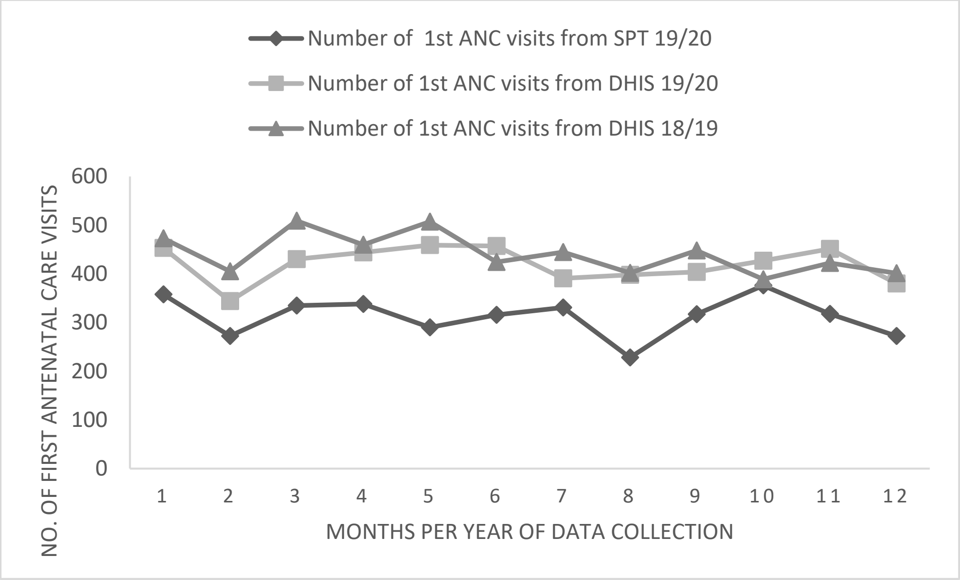


**Indicator 5a - Trends over time in client numbers for antenatal care 1st visit for SPT and DHIS 2**

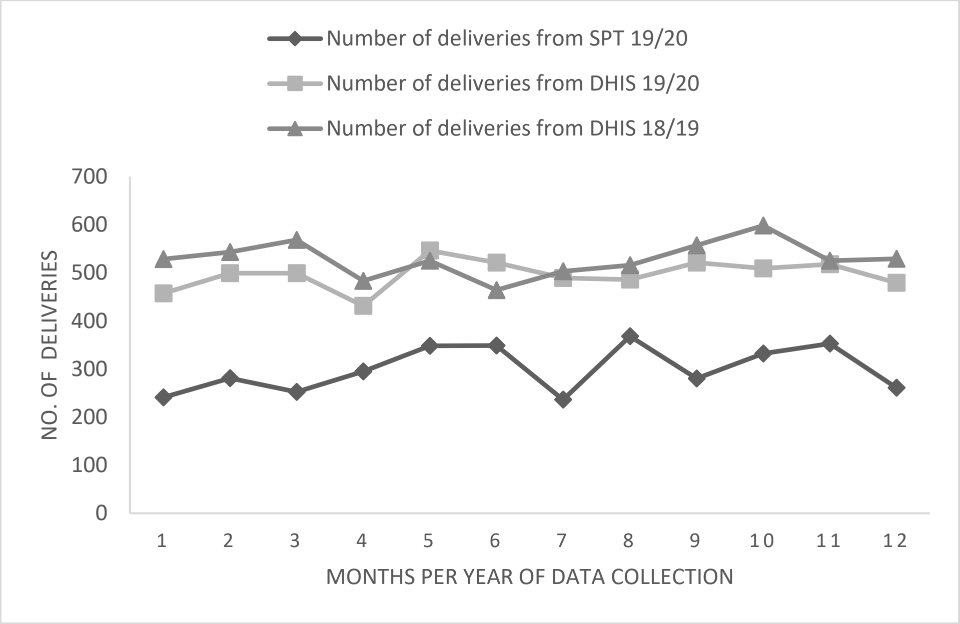


**Indicator 5b - Trends over time in number of deliveries for SPT and DHIS 2**

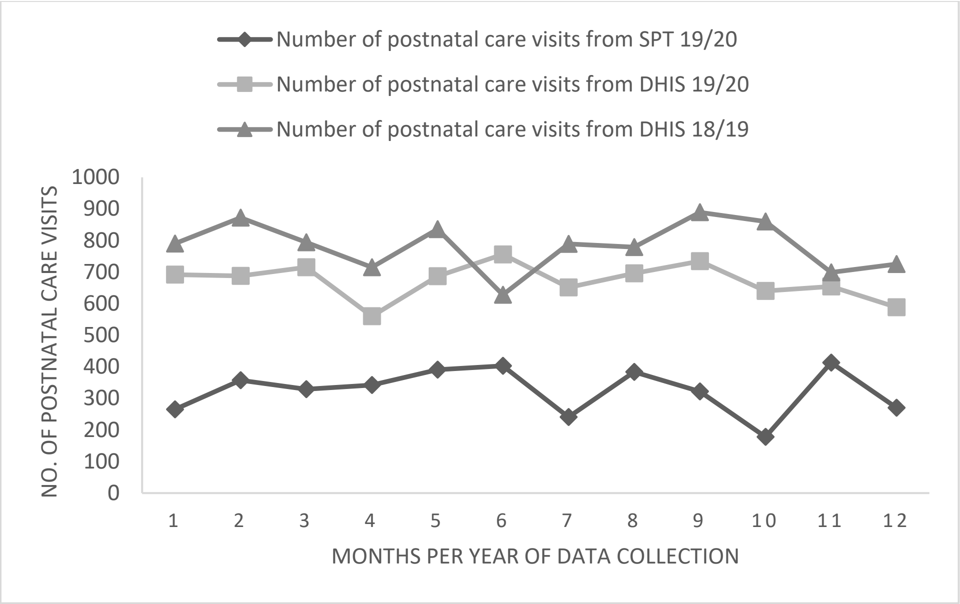


**Indicator 5c - Trends over time for number of postnatal care visits for SPT and DHIS 2**
