## Supplementary material for "Caught in the data quality trap: A case study from the evaluation of a new digital technology supporting routine health data collection in Southern Tanzania": File S1 sample SPT form

### REGISTRATION FORM

Health facility DEMO

Date (day – month)

01 1 02

Identification number **000-0092**

Name | Partner name

Age | Height (cm) | Local government chairperson

Village | Street or/Sub-village | Phone number

***Pregnancy history***

Previous CS ◯ | Age of youngest child

***Number of:***

Miscarriages/Abortions | Newborn deaths (first week)

Live births | Stillbirths | Children alive

***Current pregnancy***

Duration of pregnancy (weeks)

Date of last menstrual period (day – month)

Estimated date of delivery (day – month)

Identification number **000-0101**

Name | Partner name

Age | Height (cm) | Local government chairperson

Village | Street or/Sub-village | Phone number

***Pregnancy history***

Previous CS ◯ | Age of youngest child

***Number of:***

Miscarriages/Abortions | Newborn deaths (first week)

Live births | Stillbirths | Children alive

***Current pregnancy***

Duration of pregnancy (weeks)

Date of last menstrual period (day – month)

Estimated date of delivery (day – month)

01 1 02



DEMO

### REGISTRATION FORM

01 2 02

Identification number **000-0117**

Name ______ Partner name ______

Age ______ Height (cm) ______ Local government chairperson ______

Village ______ Street or/Sub-village ______ Phone number ______

***Pregnancy history***

Previous CS ◯ Age of youngest child ______

***Number of:***

Miscarriages/Abortions ______ Newborn deaths (first week) ______

Live births ______ Stillbirths ______ Children alive ______

***Current pregnancy***

Duration of pregnancy (weeks) ______

| | day | month |
|---|---|---|
| Date of last menstrual period | | – |
| Estimated date of delivery | | – |

Identification number **000-0125**

Name ______ Partner name ______

Age ______ Height (cm) ______ Local government chairperson ______

Village ______ Street or/Sub-village ______ Phone number ______

***Pregnancy history***

Previous CS ◯ Age of youngest child ______

***Number of:***

Miscarriages/Abortions ______ Newborn deaths (first week) ______

Live births ______ Stillbirths ______ Children alive ______

***Current pregnancy***

Duration of pregnancy (weeks) ______

| | day | month |
|---|---|---|
| Date of last menstrual period | | – |
| Estimated date of delivery | | – |

DEMO

01 2 02

### REGISTRATION UPDATE FORM

Health facility DEMO | Date day – month

02 1 02

Identification number –

Name | Partner name

Age | Height (cm) | Local government chairperson

Village | Street or/Sub-village | Phone number

***Pregnancy history***

Previous CS ◯ | Age of youngest child

***Number of:***

Miscarriages/Abortions | Newborn deaths (first week)

Live births | Stillbirths | Children alive

***Current pregnancy***

Duration of pregnancy (weeks)

Date of last menstrual period day – month

Estimated date of delivery –

Identification number –

Name | Partner name

Age | Height (cm) | Local government chairperson

Village | Street or/Sub-village | Phone number

***Pregnancy history***

Previous CS ◯ | Age of youngest child

***Number of:***

Miscarriages/Abortions | Newborn deaths (first week)

Live births | Stillbirths | Children alive

***Current pregnancy***

Duration of pregnancy (weeks)

Date of last menstrual period day – month

Estimated date of delivery –



DEMO

Shifo Karolinska Institutet IFAKARA HEALTH INSTITUTE

*Ministry of Health, Community Development, Gender, Elderly and Children, The United Republic of Tanzania*

### REGISTRATION UPDATE FORM

02 2 02

Identification number ___ – ___

Name

Partner name

Age

Height (cm)

Local government chairperson

Village

Street or/Sub-village

Phone number

***Pregnancy history***

Previous CS ◯

Age of youngest child

***Number of:***

Miscarriages/Abortions

Newborn deaths (first week)

Live births

Stillbirths

Children alive

***Current pregnancy***

Duration of pregnancy (weeks)

day month

Date of last menstrual period –

Estimated date of delivery –

Identification number ___ – ___

Name

Partner name

Age

Height (cm)

Local government chairperson

Village

Street or/Sub-village

Phone number

***Pregnancy history***

Previous CS ◯

Age of youngest child

***Number of:***

Miscarriages/Abortions

Newborn deaths (first week)

Live births

Stillbirths

Children alive

***Current pregnancy***

Duration of pregnancy (weeks)

day month

Date of last menstrual period –

Estimated date of delivery –

02 2 02



DEMO

### ANTENATAL FORM

Health facility DEMO    Date day – month

03 1 02

Identification number –

***Visit number:*** 1 ○ 2 ○ 3 ○ 4 ○ 5+ ○    Has TT card ○ TT1 ○ TT2+ ○

***Test results at current ANC visit***

BP /    Hb ,

***Urinalysis:*** Sugar ○ Protein ○ | Blood grouping and cross-match done ○

***Syphilis:*** *Woman:* Positive ○ Negative ○ Treated ○
*Partner:* Positive ○ Negative ○ Treated ○

***Other STI:*** *Woman:* Positive ○ Treated ○
*Partner:* Positive ○ Treated ○

***Repeated visits:*** No weight gain ○ Vaginal bleeding ○ Bad posture for baby ○ TB ○

***PMTCT***

| | *Woman* | *Partner* |
|---|---|---|
| Previous HIV infection | ○ | ○ |
| Received counselling before testing | ○ | ○ |
| Tested for HIV at current ANC visit | ○ | ○ |
| Tested positive | ○ | ○ |
| Counselled after testing | ○ | ○ |
| Advice on infant feeding | ○ | |
| Referred to CTC | ○ | |

***Malaria***

*mRDT/BS:* Positive ○ Negative ○ | LLIN ○ | *IPT:* 1 ○ 2 ○ 3+ ○

***Supplements provided:*** Albendazole/Mebendazole ○

Iron ○ Folic acid ○ IFA ○ Number of capsules

Referral given ○ Facility referred from

Referred to

Reason for referral

Comments

03 1 02

Shifo Karolinska Institutet IFAKARA HEALTH INSTITUTE

*Ministry of Health, Community Development, Gender, Elderly and Children, The United Republic of Tanzania*

### ANTENATAL FORM

03 2 02

Identification number ___ – ____

***Visit number:*** 1 ○ 2 ○ 3 ○ 4 ○ 5+ ○ Has TT card ○ TT1 ○ TT2+ ○

***Test results at current ANC visit***

BP ___ / ___ Hb __ , _

***Urinalysis:*** Sugar ○ Protein ○ | Blood grouping and cross-match done ○

***Syphilis:*** *Woman:* Positive ○ Negative ○ Treated ○
*Partner:* Positive ○ Negative ○ Treated ○

***Other STI:*** *Woman:* Positive ○ Treated ○
*Partner:* Positive ○ Treated ○

***Repeated visits:*** No weight gain ○ Vaginal bleeding ○ Bad posture for baby ○ TB ○

***PMTCT***

| | *Woman* | *Partner* |
|---|---|---|
| Previous HIV infection | ○ | ○ |
| Received counselling before testing | ○ | ○ |
| Tested for HIV at current ANC visit | ○ | ○ |
| Tested positive | ○ | ○ |
| Counselled after testing | ○ | ○ |
| Advice on infant feeding | ○ | |
| Referred to CTC | ○ | |

***Malaria***

*mRDT/BS:* Positive ○ Negative ○ | LLIN ○ | *IPT:* 1 ○ 2 ○ 3+ ○

***Supplements provided:*** Albendazole/Mebendazole ○

Iron ○ Folic acid ○ IFA ○ Number of capsules __

Referral given ○ Facility referred from ______

Referred to ______

Reason for referral ______

Comments ______

Health worker name ______ Signature ______

03 2 02



DEMO

### LABOUR FORM

Health facility DEMO

Date (day – month)

04 1 02

Identification number –

Name

Admission: (day – month, time)

Delivery: (day – month, time)

Labour duration over 12 hours ○

**Place of delivery**

Health facility ○ BBA ○ TBA ○ Home without TBA ○

Companion at birth ○

**Mode of delivery**

Spontaneous ○ Vacuum ○ Other ○

CS ○ Breech ○

***Cadre of birth attendant*** MO ○ AMO ○ CO/ACO ○ EN ○ RN ○ MA ○

***Birth outcome:*** Live birth ○ FSB ○ MSB ○ Multiple birth ○ *For multiple births, fill in another labour form with mother's ID and information for other newborns*

***Sex of baby:*** Female ○ Male ○

***Apgar score:*** 1 min 5 min

Breastfeeding within 1 hour ○

***Care of the baby*** No intervention ○ Stimulation ○ Bag and mask ○ Suction ○

Birth weight (g)

***Labour complications***

| | | | | | |
|---|---|---|---|---|---|
| No complications ○ | Sepsis ○ | APH ○ | Chest pain ○ | PPH ○ | Maternal exhaustion ○ |
| 3rd degree tear ○ | Anemia ○ | High blood pressure ○ | Retained placenta ○ | PROM ○ | |
| HIV (III or IV) ○ | Malaria ○ | Obstructed labour ○ | Ruptured uterus ○ | FGM ○ | |

***Before delivery*** Pre-eclampsia ○ Eclampsia ○

***During/within 24h:*** Pre-eclampsia ○ Eclampsia ○

***AMTSL:*** Oxytocine ○ Misoprostol ○ Ergometrine ○ Controlled cord traction ○ Fundal pressure ○

***EmOC:*** No intervention ○ Blood transfusion ○ Anti-hypertensive drug ○ Hysterectomy ○ Uterotonics ○

Uterine balloon tamponade ○ MVA/D&C ○ Misoprostol ○ Antibiotics ○

iv-Magnesium ○ 3rd/4th degree tear repair ○ CS/Laparotomy ○ Manual removal of placenta ○

***PMTCT:***

Previous HIV infection: Positive ○ Negative ○ Unknown ○

HIV test result during/after delivery: Positive ○ Negative ○ Unknown ○

***Discharge information***

***Feeding***

Supplements ○ Exclusive breastfeeding ○

Received ARV prophylaxis ○ Name of ARV

Maternal death: (day – month, cause of death)

Newborn death: (day – month, cause of death)

***Referrals:*** Mother ○ Baby ○ Referral for ARV at CTC ○ Referred to Referral reason

Health worker name

Signature

04 1 02

Shifo Karolinska Institutet IFAKARA HEALTH INSTITUTE

*Ministry of Health, Community Development, Gender, Elderly and Children, The United Republic of Tanzania*

DEMO

### LABOUR FORM

04 2 02

Identification number ____ – ____

Name

| | day | month | time |
|---|---|---|---|
| Admission: | | – | |
| Delivery: | | – | |

Labour duration over 12 hours ◯

**_Place of delivery_**

Health facility ◯ BBA ◯ TBA ◯ Home without TBA ◯

Companion at birth ◯

**_Mode of delivery_**

Spontaneous ◯ Vacuum ◯ Other ◯

CS ◯ Breech ◯

**_Cadre of birth attendant_** MO ◯ AMO ◯ CO/ACO ◯ EN ◯ RN ◯ MA ◯

**_Birth outcome:_** Live birth ◯ FSB ◯ MSB ◯ Multiple birth ◯ *For multiple births, fill in another labour form with mother's ID and information for other newborns*

**_Sex of baby:_** Female ◯ Male ◯ **_Apgar score:_** 1 min ___ 5 min ___ Breastfeeding within 1 hour ◯

**_Care of the baby_** No intervention ◯ Stimulation ◯ Bag and mask ◯ Suction ◯ Birth weight (g) ____

**_Matatizo na Matokeo ya Uzazi_**

No complications ◯ Sepsis ◯ APH ◯ Chest pain ◯ PPH ◯ Maternal exhaustion ◯

3rd degree tear ◯ Anemia ◯ High blood pressure ◯ Retained placenta ◯ PROM ◯

HIV (III or IV) ◯ Malaria ◯ Obstructed labour ◯ Ruptured uterus ◯ FGM ◯

**_Before delivery:_** Pre-eclampsia ◯ Eclampsia ◯

**_During/within 24h:_** Pre-eclampsia ◯ Eclampsia ◯

**_AMTSL:_** Oxytocine ◯ Misoprostol ◯ Ergometrine ◯ Controlled cord traction ◯ Fundal pressure ◯

**_EmOC:_** No intervention ◯ Blood transfusion ◯ Anti-hypertensive drug ◯ Hysterectomy ◯ Uterotonics ◯

Uterine balloon tamponade ◯ MVA/D&C ◯ Misoprostol ◯ Antibiotics ◯

iv-Magnesium ◯ 3rd/4th degree tear repair ◯ CS/Laparotomy ◯ Manual removal of placenta ◯

**_PMTCT:_**

Previous HIV infection: Positive ◯ Negative ◯ Unknown ◯

HIV test result during/after delivery: Positive ◯ Negative ◯ Unknown ◯

**_Discharge information_**

**_Feeding_**

Supplements ◯ Exclusive breastfeeding ◯

Received ARV prophylaxis ◯ Name of ARV ____

| | day | month | cause of death |
|---|---|---|---|
| Maternal death: | | – | |
| Newborn death: | | – | |

**_Referrals:_** Mother ◯ Baby ◯ Referral for ARV at CTC ◯ Referred to ____ Referral reason ____

Health worker name ____ Signature ____

04 2 02

Shifo Karolinska Institutet IFAKARA HEALTH INSTITUTE

*Ministry of Health, Community Development, Gender, Elderly and Children, The United Republic of Tanzania*

### POSTNATAL FORM

Health facility DEMO Date day __ – month __

05 1 02

Identification number ___ – ____

Maternal death ○ Newborn death ○ BP (mmHg) ___ / ___ Hb (g/dl) __ , _

**Breasts**

Normal ○

Cracked nipples ○

Mastitis ○

Abscess ○

***HIV test at postnatal ward:*** Positive ○ Negative ○

Pre-eclampsia 24h after delivery ○

Eclampsia 24h after delivery ○

***Fistula:*** Yes ○ No ○

***Mental status:*** Normal ○ Abnormal ○

***Uterus***

Normal ○ Pain ○

Not well contracted ○

***Lochia***

Normal ○

Abnormal colour ○

***Perineum:*** Healed ○ Not healed ○ Infection ○

***Family planning services provided***

Counselling ○ FP method given ○

Information materials ○ Referral for FP method ○

***Supplements given***

Iron ○ Folic acid ○ IFA ○ Number of tablets ___

Vitamin A ○ Number of tablets ___

***Mtoto***

*For multiple births, fill in another form with mother's ID and information for the baby*

Temp (°C) ___ BCG ○ OPV ○ Vitamin K ○ Weight (g) ____ Hb (g/dl) __ , _ Pallor (palms) ○ KMC initiated ○

***Infections:*** No sign of infection ○ Umbilical cord ○ Skin ○ Oral ○ Eye ○ Jaundice ○ Sepsis ○

***PMTCT:*** *HIV test result:* Positive ○ Negative ○ ARV given ○ Name of ARV ___ Duration (weeks) ___

***Feeding:*** Exclusive ○ Replacement ○ Mixed ○

***Rufaa***

Referral from ___ Referral to ___ Reason ___

05 1 02

### POSTNATAL FORM

05 2 02

Identification number ___ – ___

Maternal death ○ Newborn death ○ BP (mmHg) ___ / ___ Hb (g/dl) ___,___

***HIV test at postnatal ward:*** Positive ○ Negative ○

Pre-eclampsia 24h after delivery ○

Eclampsia 24h after delivery ○

***Fistula:*** Yes ○ No ○

***Mental status:*** Normal ○ Abnormal ○

***Uterus***

Normal ○ Pain ○

Not well contracted ○

***Lochia***

Normal ○

Abnormal colour ○

***Breasts***

Normal ○

Cracked nipples ○

Mastitis ○

Abscess ○

***Perineum:*** Healed ○ Not healed ○ Infection ○

***Family planning services provided***

Counselling ○ FP method given ○

Information materials ○ Referral for FP method ○

***Supplements given***

Iron ○ Folic acid ○ IFA ○ Number of tablets ___

Vitamin A ○ Number of tablets ___

***Mtoto***

*For multiple births, fill in another form with mother's ID and information for the baby*

Temp (°C) ___ BCG ○ OPV ○ Vitamin K ○ Weight (g) ___ Hb (g/dl) ___,___ Pallor (palms) ○ KMC initiated ○

***Infections:*** No sign of infection ○ Umbilical cord ○ Skin ○ Oral ○ Eye ○ Jaundice ○ Sepsis ○

***PMTCT:*** *HIV test result:* Positive ○ Negative ○ ARV given ○ Name of ARV ___ Duration (weeks) ___

***Feeding:*** Exclusive ○ Replacement ○ Mixed ○

***Rufaa***

Referral from ___ Referral to ___ Reason ___

Health worker name ___ Signature ___

05 2 02

Shifo Karolinska Institutet IFAKARA HEALTH INSTITUTE

*Ministry of Health, Community Development, Gender, Elderly and Children, The United Republic of Tanzania*
