## Supplementary material for "Caught in the data quality trap: A case study from the evaluation of a new digital technology supporting routine health data collection in Southern Tanzania": Table S1 Glossary of terms

| | |
|---|---|
| Health Information System (HIS) | An "*integrated effort to collect, process, report and use health information and knowledge to influence policy-making, programme action and research*". |
| Health Management Information System (HMIS) | "*A system of service-generated data derived from facilities and patient–provider interactions covering aspects such as care offered, quality of care and treatments administered*" as part of the HIS. |
| District Health Information System 2 (DHIS2) | The **web-based** software platform to collect, access and analyze data as the electronic part of HMIS in Tanzania. |
| Facility registers | **Paper-based** registers as part of the HMIS for manual data collection on service provision for individual clients in all departments of a health facility including maternal health care. Individual data is summarized daily in paper-based tally sheets. |
| Facility summary reports | **Paper-based** forms as part of the HMIS to summarize monthly data on each client per department including maternal health care. |
| SPT forms | **Paper-based** forms as part of SPT system for manual data collection on service provision for individual clients using a unique identifier. One form is completed for each client-provider encounter using the same identifier for an individual client. The **equivalent for HMIS** at facility level is the individual data entry in a row of the **facility register**. |
| Electronic SPT summary reports | Electronic monthly summary of SPT data collected from each facility accessible on the SPT dashboard. The **equivalent for HMIS** is the electronic report for each facility in **DHIS2**. |
