## Supplementary material for "Caught in the data quality trap: A case study from the evaluation of a new digital technology supporting routine health data collection in Southern Tanzania": Table S2 Indicator 3

**Table S2 Indicator 3: Reporting completeness for ANC 1st visit for DHIS2 and SPT by facility**

| Facility ID | DHIS2 | | SPT 2019/2020 |
|---|---|---|---|
| | 2018/2019 | 2019/2020 | |
| | *Total number of months with non-missing reporting on ANC 1st visit (%)* | | |
| 1 HOSP | 12 (100) | 12 (100) | 12 (100) |
| 2 HC | 12 (100) | 12 (100) | 12 (100) |
| 3 HC | 12 (100) | 12 (100) | 12 (100) |
| 4 D | 12 (100) | 12 (100) | 12 (100) |
| 5 D | 12 (100) | 11 (92) | 11 (92) |
| 6 D | 12 (100) | 12 (100) | 10 (83) |
| 7 D | 12 (100) | 12 (100) | 12 (100) |
| 8 D | 12 (100) | 12 (100) | 12 (100) |
| 9 D | 10 (83) | 11 (92) | 10 (83) |
| 10 D | 11 (92) | 10 (83) | 12 (100) |
| 11 D | 12 (100) | 11 (92) | 6 (50) |
| 12 D | 11 (92) | 12 (100) | 11 (92) |
| 13 D | 11 (92) | 12 (100) | 12 (100) |

*Legend: HOSP= Hospital, HC= Health Center, D= Dispensary, Grey= > 90%*
