## Supplementary material for "Caught in the data quality trap: A case study from the evaluation of a new digital technology supporting routine health data collection in Southern Tanzania": Table S3 Indicator 7

**Table S3: Indicator 7- Difference between number of deliveries and women receiving oxytocin after birth of < +/- 10 % in DHIS2 and SPT by facility**

| Facility ID | DHIS2 | | | | | | SPT 2019/2020 | | |
|---|---|---|---|---|---|---|---|---|---|
| | 2018/2019 | | | 2019/2020 | | | | | |
| | *No. of deliveries* | *Oxytocin given* | *Ratio* | *No. of deliveries* | *Oxytocin given* | *Ratio* | *No. of deliveries* | *Oxytocin given* | *Ratio* |
| 1 HOSP | 3,214 | 3,129 | 0.97 | 2.904 | 2,527 | 0.87 | 2,105 | 1,916 | 0.91 |
| 2 HC | 613 | 550 | 0.90 | 608 | 593 | 0.98 | 375 | 338 | 0.90 |
| 3 HC | 1,230 | 1,066 | 0.87 | 1,249 | 1,025 | 0.82 | 448 | 385 | 0.86 |
| 4 D | 80 | 75 | 0.94 | 77 | 69 | 0.90 | 50 | 47 | 0.94 |
| 5 D | 140 | 136 | 0.97 | 123 | 106 | 0.86 | 69 | 64 | 0.93 |
| 6 D | 187 | 171 | 0.91 | 155 | 142 | 0.92 | 119 | 104 | 0.87 |
| 7 D | 359 | 356 | 0.99 | 366 | 356 | 0.97 | 217 | 187 | 0.86 |
| 8 D | 43 | 35 | 0.81 | 32 | 24 | 0.75 | 18 | 14 | 0.78 |
| 9 D | 139 | 128 | 0.92 | 174 | 174 | 1.00 | 88 | 81 | 0.92 |
| 10 D | 102 | 93 | 0.91 | 95 | 74 | 0.78 | 21 | 21 | 1.00 |
| 11 D | 65 | 57 | 0.88 | 21 | 20 | 0.95 | 2 | 2 | 1.00 |
| 12 D | 52 | 42 | 0.81 | 36 | 32 | 0.89 | 16 | 16 | 1.00 |
| 13 D | 115 | 88 | 0.77 | 115 | 91 | 0.79 | 68 | 62 | 0.91 |

*Legend: HOSP= Hospital, HC= Health Center, D= Dispensary, Grey= Difference < +/- 0.10*
